# Associations of menopause hormone therapy with late-life cognition and brain health vary by formulation and *APOEε4* presence

**DOI:** 10.64898/2026.09.16.26363120

**Authors:** Mateja Perović, Laura L. Gravelsins, Madeline Wood Alexander, Andrew J. McGovern, Kelly J. Murphy, Jennifer S. Rabin, Liisa A.M. Galea

**Author notes:** Corresponding authors: Liisa Galea, PhD, Centre for Addiction and Mental Health, Mateja Perović, PhD, Centre for Addiction and Mental Health.

## Abstract

Implications of menopause hormone therapy (MHT) for cognitive aging remain unclear despite decades of research. Potential reasons include insufficient consideration of MHT type, despite notable pharmacokinetic differences across formulations, and limited attention to individual differences that may exacerbate cognitive risk. We examined late-life neurocognitive outcomes as a function of *APOEε4* status, the strongest genetic predictor of Alzheimer’s disease, and MHT formulation (N=8,741). Results suggest formulation- and domain-specific association between MHT and cognition, with evidence of estradiol-based MHT providing more benefits for *APOEε4* carriers relative to conjugated equine estrogens use. Neuroimaging analyses available for a subset of participants (N=930) revealed a consistent pattern of more favourable associations (higher volume, cortical thickness; less white matter damage) in *APOEε4* carriers taking estradiol-based MHT. These results reframe mixed MHT findings as an issue of exposure definition and support formulation- and genetic-risk-aware approaches to menopause care.

## INTRODUCTION

Most people with Alzheimer’s disease (AD) are women^1^ but the reasons for this disparity are still being elucidated. Critically, AD pathogenesis begins as early as midlife, decades before symptom onset^2,3^. For most women, midlife coincides with the menopausal transition. This period of declining ovarian function has been identified as a neurological transition state^4^, an inflection point for cognitive aging^5^, and a potential contributing factor to the greater prevalence of AD in women^6,7^. The menopausal transition is associated with a range of changes in brain structure, function, and metabolism^8^. Similarly, memory score declines have been reported during and after the menopausal transition, along with alterations in associated neurocircuitry^6,9^. Perhaps unsurprisingly, around 60% of individuals report subjective cognitive difficulties that affect their daily life during the menopausal transition^10^.

A likely mechanism underlying many of the menopause-associated cognitive and brain changes is the loss of ovarian hormones that can support cognition. Estrogens, such as estradiol (E2; the most bioactive estrogen in humans) and estrone (E1), and progesterone decline across the menopausal transition. E2 is a powerful neuromodulator with significant effects on brain structure^11,12^, function^13,14^, plasticity^15^ and cognitive performance^12,16^. The decline of E2 across the menopausal transition has been linked with reduced memory scores^6,13^. E1, which also declines with menopause but to a lesser extent than E2, is a less potent estrogen with fewer positive effects on cognition and neuroplasticity in animal studies^17^. Particularly strong evidence for the importance of ovarian hormones for cognition comes from the literature on surgical menopause, which involves an abrupt loss of ovarian hormones due to surgical removal of the ovaries. Surgical menopause, particularly prior to the age of spontaneous menopause, is associated with increased risk of dementia^18^, lower medial temporal lobe volumes^19,20^, increased neuropathology^21^, and lower episodic memory scores^19,20^, findings which are largely attributed to abrupt ovarian hormone loss following surgery.

The significant associations between ovarian hormones and cognition across the menopausal transition have prompted work examining menopausal hormone therapy (MHT) as a potential pharmacological intervention for mitigating negative cognitive outcomes and reducing risk of Alzheimer’s disease^22^. However, this literature has yielded highly mixed findings^22–24^. This may be in part due to a lack of attention paid to hormonal formulation of MHT^23,25^. Even though commonly studied forms of MHT differ in pharmacodynamics and receptor affinities, most studies in the literature do not differentiate between different types of MHT (e.g., characteristics summarized in a recent meta-analysis^26^). In particular, conjugated equine estrogens (CEE) are comprised largely of E1-sulphate which has a lower affinity for binding to estrogen receptors in the brain compared to E2^23^. On the other hand, E2-based MHT should have more potent effects on the brain which may depend on route of administration^23,25^. Although oral E2 largely converts to E1, depending on conjugation of E2, transdermal and vaginal E2 bypass first-pass metabolism by the liver resulting in higher E2 availability^27^. Use of concurrent progesterone or progestins, required to protect the endometrium from neoplasms post-menopause, may also affect cognitive outcomes^28^. The addition of medroxyprogesterone acetate (MPA) to estrogens-based therapies has been associated with negative cognitive outcomes, but this has not been the case for micronized progesterone^29,30^. Thus, the composition of MHT is critical to consider when assessing MHT effects on cognition.

Another important factor is the presence of individual risk factors for cognitive decline. Notably, the presence of apolipoprotein E ε4 allele (*APOEε4*) is the strongest genetic predictor of sporadic AD onset and progression. Moreover, female carriers face higher risk of AD relative to male carriers, particularly after the age of menopause^1,31,32^. Cognitively healthy *APOEε4* female carriers have also been shown to have lower episodic memory performance^33^, hippocampal connectivity^34^, and default mode network activity^35^ relative to male carriers. Limited evidence suggests that the use of estrogen MHT (unspecified type) may decrease risk of AD in female *APOEε4* carriers^36^ and that MHT (unspecified formulation) is associated with higher volumes of medial temporal lobe regions^37,38^ which are among the first areas to atrophy in AD^39^. These findings underscore the importance of considering *APOE* genotype as a potential modulator of MHT effects on cognition and brain health.

The current study examined the interacting associations of MHT and *APOEε4* on late-life cognition across a range of cognitive domains, while taking specific hormonal formulation of MHT into account. Baseline behavioral outcomes were predicted in cognitively healthy participants from the National Alzheimer’s Coordinating Centre (NACC) either not taking MHT or taking one of the four most commonly studied MHT formulations: E2 only, E2+P (progesterone or progestin, excluding MPA), CEE only, or CEE+MPA. As there are significant disparities in menopausal care in the United States^40^, and experiences of menopause itself, such as timing and symptom burden, differ by race among American women^41^, we conducted exploratory analyses including race as a potential modifier of associations between MHT and cognition. Beyond cross-sectional comparisons, cognitive trajectories before and after MHT discontinuation were assessed by APOEε4 status. Finally, structural brain outcomes were predicted in a subset of participants with available scan data. Results indicate that while both E2 and CEE show some cognitive benefits, they vary by cognitive domain and APOEε4 status. E2 in particular shows more promise for cognitive and brain health in APOEε4 carriers.

## RESULTS

### Cognitive performance

#### Cognitive outcomes differed by domain and MHT formulation

Cognitive outcomes of interest included episodic memory, verbal performance (category fluency, confrontation naming), and executive function – cognitive domains previously shown to vary across the menopausal transition and as a function of MHT use^24,42,43^ (Figure 1A; demographic characteristics in Table 1). Results (Figure 1B) indicated that associations between MHT and cognition varied by MHT formulation and cognitive domain. Participants taking CEE had lower episodic memory scores than participants taking E2 (*p*=0.006, 95% CI [0.36,2.17]) as well as participants taking no MHT (*p*=0.017, 95% CI [0.15,1.57]), but CEE showed some benefits for verbal performance as participants taking CEE had higher category fluency scores than both participants not taking MHT (*p*=0.037, 95% CI [0.04,1.17]) and participants taking E2+P (*p*=0.028, 95% CI [0.15,2.74]). Both participants taking CEE (*p*=0.018, 95% CI [−2.25,−0.21]) and participants taking E2 (*p*=0.033, 95% CI [−2.06,−0.09]) had higher naming task scores than participants taking CEE+MPA. All significant results with the exception of the E2-CEE+MPA association for the naming task were robust to bootstrap resampling (p_boot_<0.05; N_boot_=1,000).

**Figure 1.**
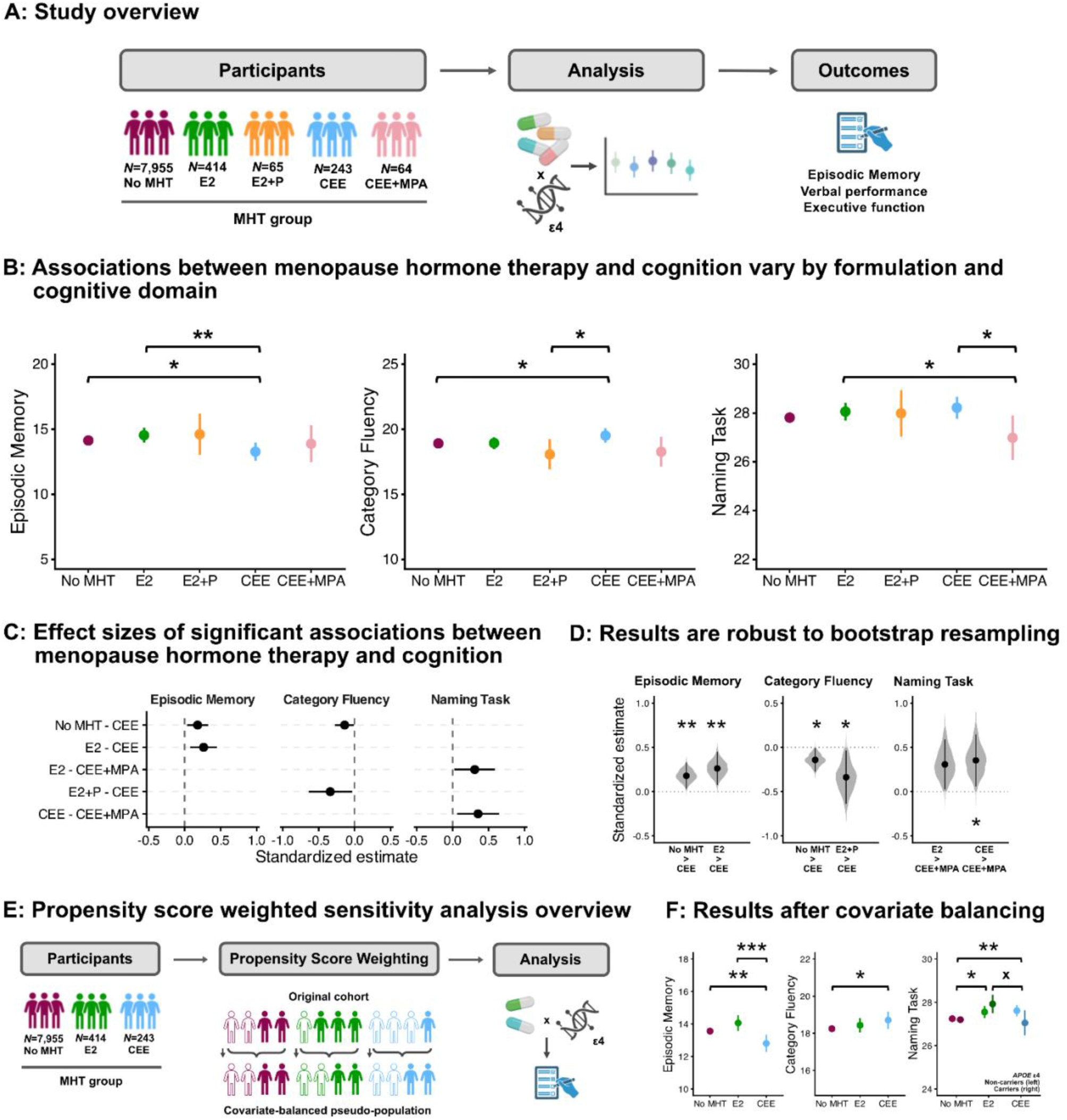
Associations between menopause hormone therapy formulation and cognitive performance. A: Cognitive outcomes were predicted as a function of an interaction between menopause hormone therapy (MHT) use and *APOEε4* status. MHT groups included participants either taking no MHT (n=7,955), or one of the four most commonly studied MHT formulations: estradiol (E2; n=414), combination estradiol and progestogens (E2+P; n=65), conjugated equine estrogens (CEE; n=243), or CEE and medroxyprogesterone acetate (CEE+MPA; n=64). B: Associations between menopause hormone therapy and cognitive performance vary by MHT formulation and cognitive domain. Participants taking CEE had worse episodic memory performance than participants not taking menopause hormone therapy and participants taking E2. Participants taking CEE had better category fluency performance relative to participants not taking MHT and those taking E2+P. Participants taking E2 and participants taking CEE had better naming task performance than participants taking CEE+MPA. Models were adjusted for age, years of education, depressive symptoms, cardiovascular risk, and race/ethnicity. Plots show model estimates with error bars denoting 95% confidence intervals. There were no significant interactions between MHT and *APOEε4* status, but a breakdown of model estimates by *APOEε4* status is provided for descriptive purposes in Extended Figure 1. C: Standardized effect sizes of significant effects in primary analytic models, with values expressed in standard deviation units. D: Results of 1,000 bootstrap resamples per outcome suggest that significant results are mostly robust to resampling with replacement. Points and bars represent bootstrapped standardized estimates with 95% confidence intervals. Grey violins represent the bootstrap sampling distribution of the effect estimate across bootstraps. D: Primary analyses were followed by propensity score weighted sensitivity analyses in a subset of participants (no MHT, E2, CEE) to improve covariate balance and help account for potential selection bias in the MHT groups. E: Results of the propensity score weighted sensitivity analyses were convergent with those of the primary analyses. Both participants taking E2 and those not taking MHT outperformed participants taking CEE on episodic memory. Participants taking CEE had better category fluency performance relative to participants not taking MHT. Both participants taking E2 and participants taking CEE had better performance relative to participants not taking MHT on the naming task. Additionally, propensity score weighted analyses of the naming task performance yielded a significant MHT by APOEε4 interaction for the E2 and CEE groups, suggesting more beneficial associations for carriers in the E2 group and non-carriers in the CEE group. Models were adjusted for age, years of education, depressive symptoms, cardiovascular risk, and race/ethnicity. Plots show model estimates with error bars denoting 95% confidence intervals. For all plots, “*” indicates p < .05, “**” indicates p < .01, “***” indicates p < .001, “x” indicates a significant interaction. The study overview figure was created using BioRender.

**Table 1.**
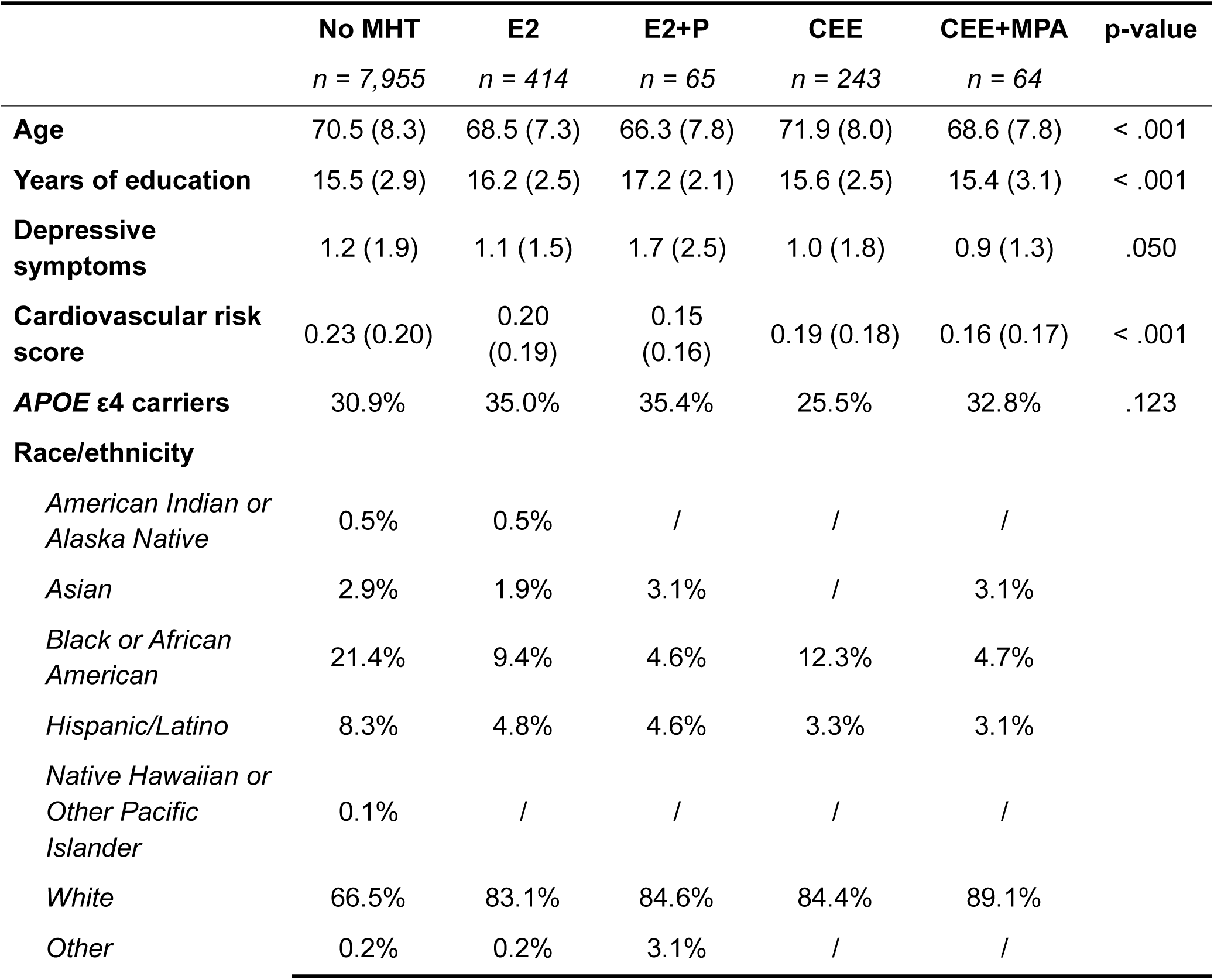
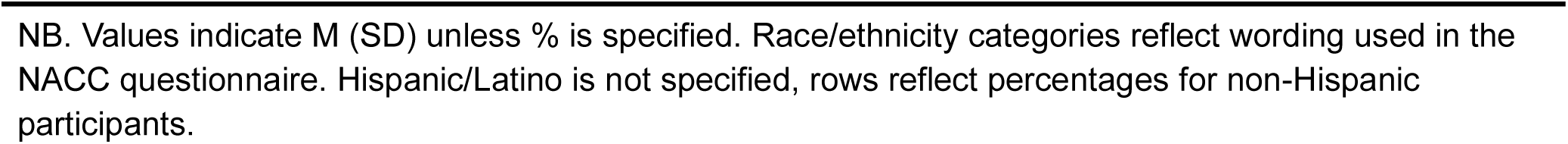
Cognitive analyses cohort characteristics at baseline.

|  | No MHT<br>*n* = 7,955 | E2<br>*n* = 414 | E2+P<br>*n* = 65 | CEE<br>*n* = 243 | CEE+MPA<br>*n* = 64 | p-value |
| --- | --- | --- | --- | --- | --- | --- |
| **Age** | 70.5 (8.3) | 68.5 (7.3) | 66.3 (7.8) | 71.9 (8.0) | 68.6 (7.8) | < .001 |
| **Years of education** | 15.5 (2.9) | 16.2 (2.5) | 17.2 (2.1) | 15.6 (2.5) | 15.4 (3.1) | < .001 |
| **Depressive symptoms** | 1.2 (1.9) | 1.1 (1.5) | 1.7 (2.5) | 1.0 (1.8) | 0.9 (1.3) | .050 |
| **Cardiovascular risk score** | 0.23 (0.20) | 0.20 (0.19) | 0.15 (0.16) | 0.19 (0.18) | 0.16 (0.17) | < .001 |
| **APOE ε4 carriers** | 30.9% | 35.0% | 35.4% | 25.5% | 32.8% | .123 |
| **Race/ethnicity** |  |  |  |  |  |  |
| *American Indian or Alaska Native* | 0.5% | 0.5% | / | / | / |  |
| *Asian* | 2.9% | 1.9% | 3.1% | / | 3.1% |  |
| *Black or African American* | 21.4% | 9.4% | 4.6% | 12.3% | 4.7% |  |
| *Hispanic/Latino* | 8.3% | 4.8% | 4.6% | 3.3% | 3.1% |  |
| *Native Hawaiian or Other Pacific Islander* | 0.1% | / | / | / | / |  |
| *White* | 66.5% | 83.1% | 84.6% | 84.4% | 89.1% |  |
| *Other* | 0.2% | 0.2% | 3.1% | / | / |  |

NB. Values indicate M (SD) unless % is specified. Race/ethnicity categories reflect wording used in the NACC questionnaire. Hispanic/Latino is not specified, rows reflect percentages for non-Hispanic participants.

As this was an observational study, the primary analyses were followed by propensity score weighted sensitivity analyses to account for potential selection bias in participants taking MHT (Figure 1E). Due to the low sample sizes following weighting in the combined (E2+P; CEE+MPA) groups, the sensitivity analyses were conducted using data from participants taking E2, CEE, or not taking MHT. All primary results remained significant (Figure 1). Both participants taking E2 (*p*<0.001, 95% CI [-2.42, -0.68]) and those not taking MHT (*p*=0.005, 95% CI [-1.61, -0.29]) outperformed participants taking CEE. Additionally, a marginally significant association indicating better performance in participants taking E2 relative to those not taking MHT emerged following covariate balancing (*p*=0.051, 95% CI [-0.00, 1.20]). Significant results regarding verbal performance in the propensity weighted analyses indicated better performance in participants taking CEE relative to those not taking MHT for category fluency (*p*=0.012, 95% CI [0.15, 1.25]), and both participants taking E2 (*p*=0.025, 95% CI [0.04, 0.58]) and participants taking CEE (*p*=0.006, 95% CI [0.11, 0.64]) relative to those not taking MHT for the naming task. Additionally, propensity score weighted analyses of the naming task performance yielded a significant MHT by APOEε4 interaction (*p*=0.023, 95% CI [-1.75, - 0.13]) for the E2 and CEE groups, suggesting more benefits to carriers in the E2 group and non-carriers in the CEE group.

#### Menopause hormone therapy showed more beneficial associations in Black participants

Exploratory analyses examined cognitive outcomes as a function of MHT use and race in participants using E2, CEE, or no MHT (Figure 2A; demographic characteristics in Table 2). Results (Figure 2B) suggested a different pattern of associations between MHT and cognitive outcomes in Black participants compared to the overall sample analysis (Figure 2). Using E2 was associated with better naming task performance relative to not using MHT in Black participants (*p*=0.008, CI[0.32,2.17]) with a significant ethnicity ^x^ MHT interaction indicating that this differed from the pattern seen in white participants (*p*=0.048, CI[-1.20,0.01]). Similarly, both E2 (*p*=0.046, CI[0.35,36.14]) and CEE (*p*=0.030, CI[1.53,29.41]) were associated with faster Trail Making Test B completion relative to no MHT in Black participants. The latter differed from the direction of association in white participants (*p*=0.002, CI[-37.85,-8.74]). Finally, better episodic memory in E2 relative to CEE users, an association seen in the pooled cohort, remained significant in Black participants (*p*=0.047, CI[-4.39,-0.03]) but this was not the case for the positive association between CEE use and category fluency seen in the pooled cohort.

**Figure 2.**
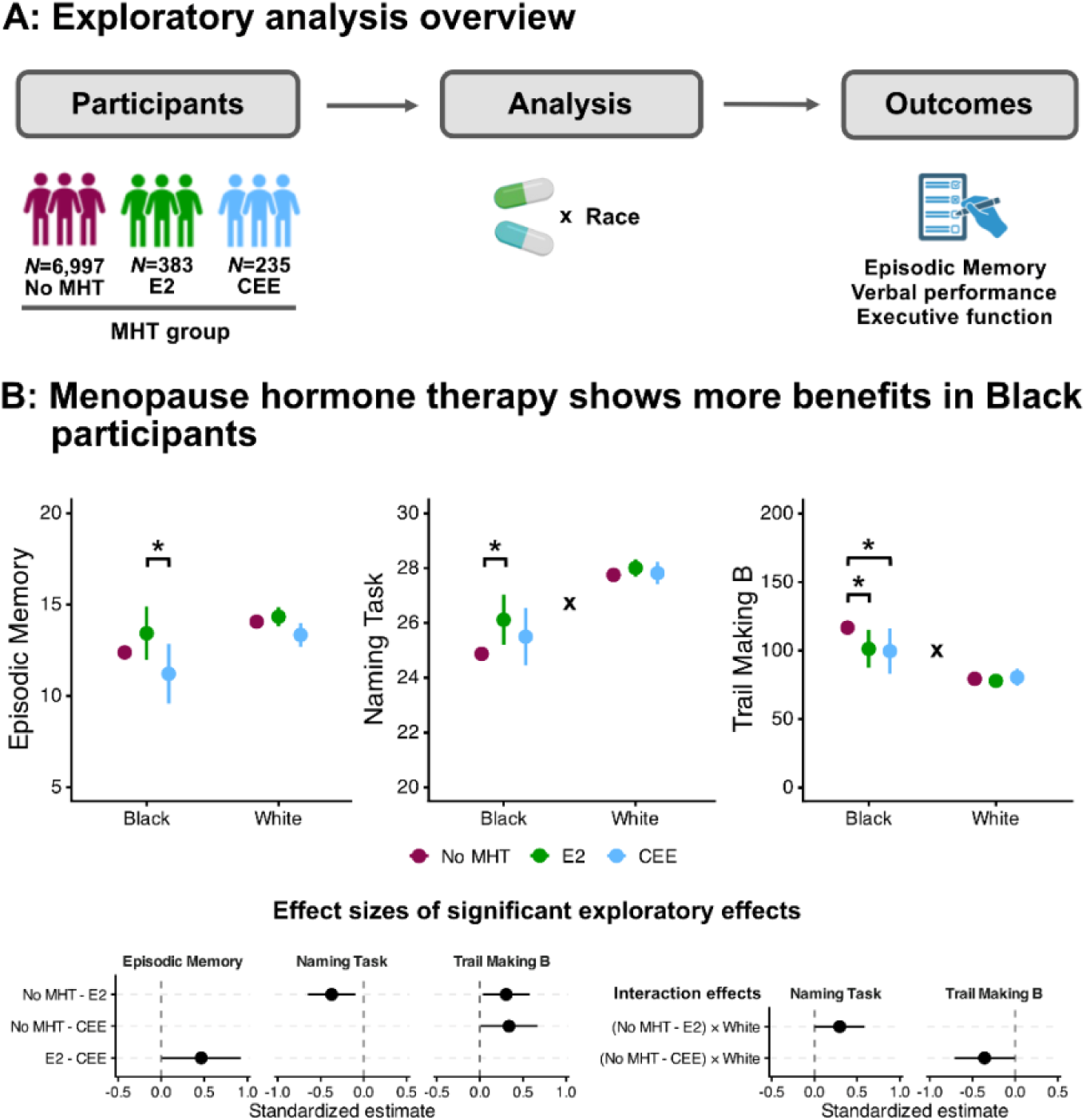
Associations between menopause hormone therapy formulations and cognitions vary by race. A: Exploratory analyses examined cognitive outcomes as a function of menopause hormone therapy (MHT) use and race in a sample of Black and white participants who either did not use MHT (N = 6,997) or used estradiol (E2; N = 383) or conjugated equine estrogens (CEE; N = 235). All exploratory analyses were conducted with Black participants as the reference group. Main effects of MHT in these analyses can thus be interpreted as effects in Black participants, with significant interactions indicating difference in direction of effect relative to white participants. B: Results indicated increased more beneficial associations between menopause hormone therapy use and cognitive performance for Black participants. E2 use was associated with better episodic memory performance in Black participants, relative to taking CEE. E2 was associated with better naming task performance relative to not taking MHT in Black but not white participants. Both E2 and CEE use was associated with faster Trail Making Test B completion times in Black but not white participants. Models were adjusted for age, years of education, cardiovascular risk, and *APOEε4* status. Plots show model estimates with error bars denoting 95% confidence intervals. “*” indicates p < .05, “**” indicates p < .01, “***” indicates p < .001, “x” indicates a significant interaction. Standardized effect sizes of significant main (left) and interaction (right) effects are shown with values expressed in standard deviation units. The analysis overview figure was created using BioRender.

**Table 2.** Demographic characteristics by ethnicity for exploratory analyses.

|  | No MHT |  | E2 |  | CEE |  |
| --- | --- | --- | --- | --- | --- | --- |
|  | White<br>*n* = 5,291 | Black<br>*n* = 1,706 | White<br>*n* = 344 | Black<br>*n* = 39 | White<br>*n* = 205 | Black<br>*n* = 30 |
| **Age** | 70.9 (8.6) | 70.4 (7.4) | 68.6 (7.4) | 68.1 (5.4) | 72.5 (8.1) | 68.9 (6.1) |
| **Years of education** | 15.9 (2.5) | 15.0 (2.8) | 16.4 (2.4) | 15.9 (2.5) | 15.7 (2.5) | 15.2 (2.3) |
| **Depressive symptoms** | 1.2 (1.8) | 1.1 (1.7) | 1.1 (1.5) | 1.2 (1.8) | 1.0 (1.8) | 1.0 (1.2) |
| **Cardiovascular risk score** | 0.20<br>(0.19) | 0.32<br>(0.21) | 0.17<br>(0.17) | 0.32<br>(0.24) | 0.18<br>(0.17) | 0.30<br>(0.21) |
| **APOE ε4 carriers** | 30.3% | 36.7% | 35.2% | 38.5% | 23.9% | 30.0% |

NB. Values indicate mean (SD) unless % is specified.

#### Cognitive trajectories differed before and after MHT discontinuation

Trajectories pre- and post-MHT (E2, CEE) discontinuation were analyzed in a subset of participants with at least two consecutive visits during which MHT use was reported (pre-discontinuation) followed by at least two consecutive visits during which no MHT use was reported (post-discontinuation)(Figure 2A). Demographic characteristics of participants in the E2 and CEE within-subject samples are presented in Table 3.

**Table 3.**
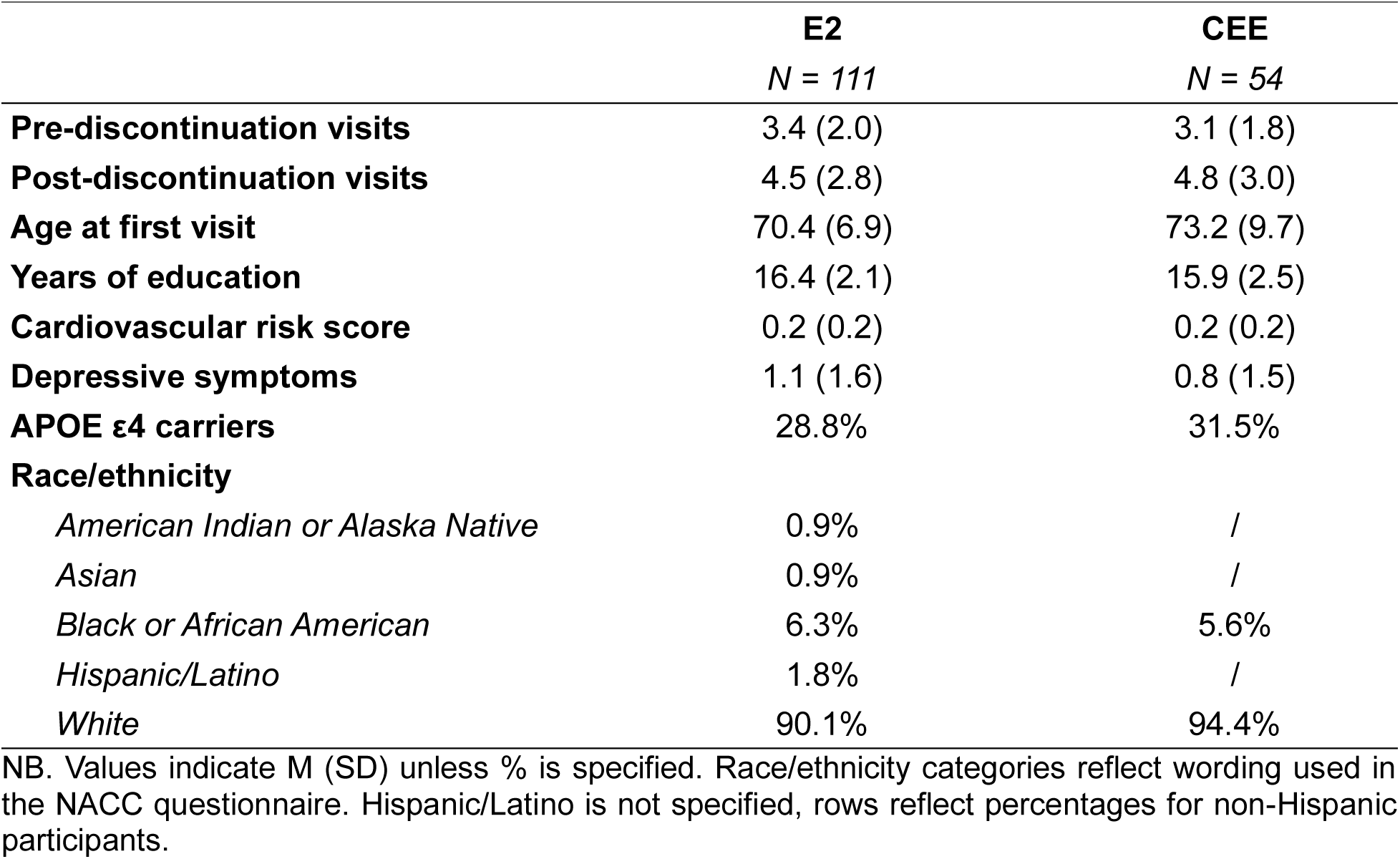
Demographic characteristics of the pre-/post-discontinuation analysis samples.

Among E2 users (Figure 3B; left), episodic memory performance remained stable both pre- and post-discontinuation (slopes *p*>0.05). For category fluency, pre-discontinuation trajectories were flat (*p* > 0.05) but both *APOEε4* non-carriers (p=0.006, 95% CI [-0.41,-0.07]) and carriers (p=0.025, 95% CI [-0.45,-0.03]) showed negative trajectories post-discontinuation. This change in slope directions following discontinuation was significant for non-carriers (*p*=0.007, 95% CI [-0.57,-0.09]). For the naming task, *APOEε4* carriers had a positive pre- (*p*<0.002, 95% CI [0.09,0.38]) but not post-discontinuation slope, a significant change in trajectory (*p*=0.002, 95% CI [-0.43,-0.10]). Finally, positive pre- (*p*=0.012, 95% CI [0.42,3.36]) and post-discontinuation (*p*=0.001, 95% CI [1.39,4.98]) slopes in non-carriers for Trail Making Test B indicated increasingly slower task completion over time, consistent with gradual age-related decline.

**Figure 3.**
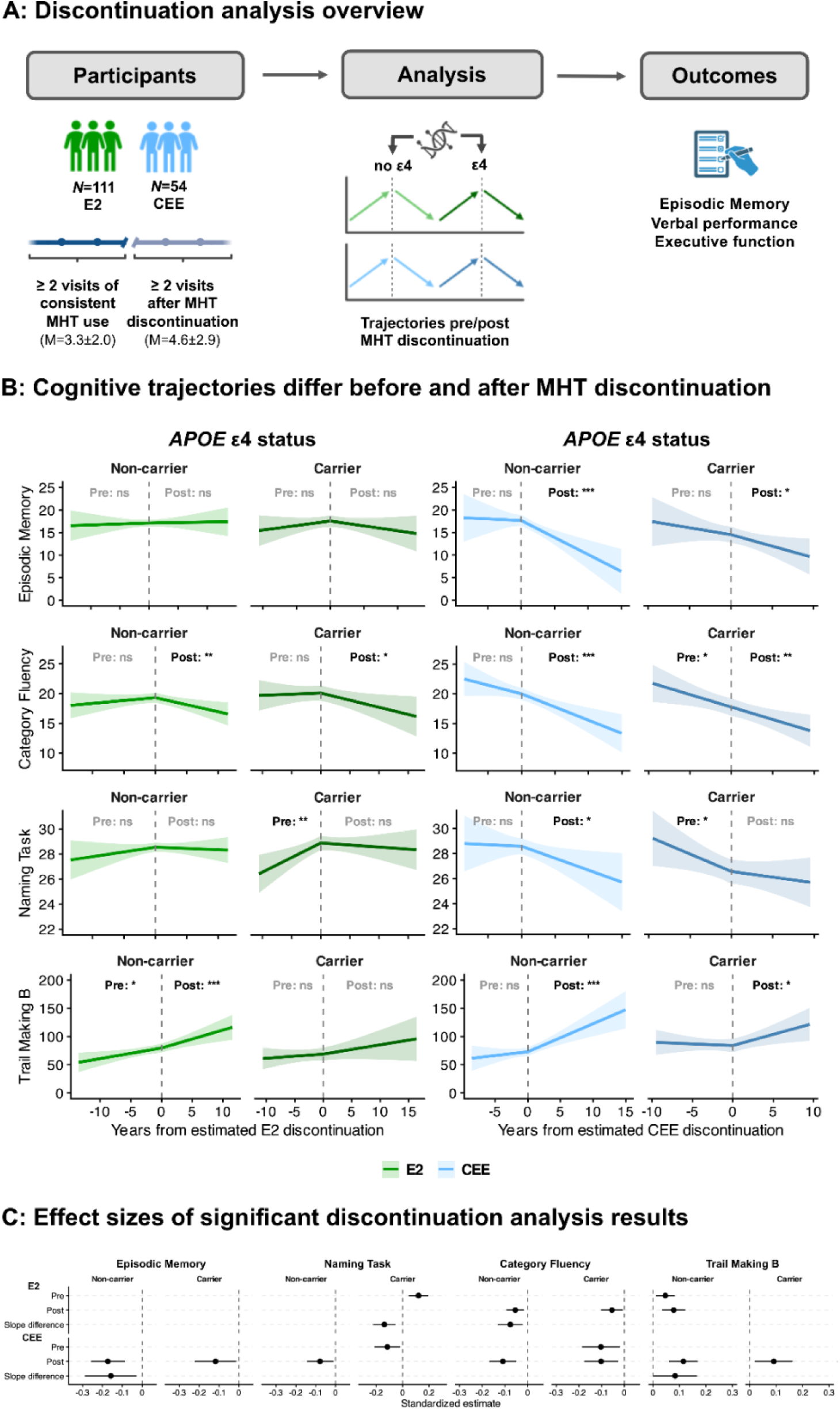
Cognitive trajectories before and after menopause hormone therapy discontinuation. A: Trajectories before and after MHT discontinuation were analyzed in a subset of participants (N = 165) with at least two consecutive visits during which MHT use was reported (pre-discontinuation) followed by at least two consecutive visits during which no MHT use was reported (post-discontinuation). Discontinuation was estimated at the midpoint between the pre- and post-discontinuation visits. On average, participants had 3.3 ± 2 pre-discontinuation visits and 4.6 ± 2.9 post-discontinuation visits. Modeling was performed in two separate within-subject samples, one of E2 (N = 111) and one of CEE (N = 54) users. B: Among E2 users, both *APOEε4* non-carriers and carriers showed negative post-discontinuation trajectories for category fluency; *APOEε4* carriers had a positive pre-but not post-discontinuation slope for the naming task; and *APOEε4* non-carriers had positive pre- and post-discontinuation slopes for Trail Making Test B, indicated increasingly slower task completion over time. Among CEE users, both *APOEε4* non-carriers and carriers showed negative episodic memory slopes post-discontinuation; *APOEε4* non-carriers showed a negative slope post-discontinuation whereas carriers had negative slopes both before and after discontinuation for category fluency; *APOEε4* non-carriers had negative trajectories post-discontinuation, whereas carriers had negative trajectories pre-discontinuation for the naming task; and both *APOEε4* non-carriers and carriers showed positive slopes, indicating slower performance, post-discontinuation for Trail Making Task B. All models were adjusted for age at baseline, years of education, race/ethnicity, depressive symptoms, cardiovascular risk, and the cumulative proportion of visits for which participants had reported MHT use. Square root of visit number was added as a covariate to models predicting episodic memory and naming task performance as visual inspection suggested presence of practice effects (Extended Figure 2). Plots show model estimates with shaded regions denoting 95% confidence intervals. “*” indicates p < .05, “**” indicates p < .01, “***” indicates p < .001, “x” indicates a significant interaction. C: Standardized effect sizes of significant discontinuation analysis effects, with values expressed in standard deviation units. The analysis overview figure was created using BioRender.

Among CEE users (Figure 3B; right), both *APOEε4* non-carriers (*p*<0.001, 95% CI [-1.12,-0.38]) and carriers (*p*=0.028, 95% CI [-0.97,-0.06]) showed negative episodic memory slopes post-discontinuation, a significant change in trajectory for non-carriers (*p*=0.018, 95% CI [-1.25,-0.12]). For category fluency, *APOEε4* non-carriers showed a negative slope post-discontinuation (*p*=0.001, 95% CI [-0.69,-0.20]) whereas carriers had negative slopes both before (*p*=0.017, 95% CI [-0.76,-0.08]) and after (*p*=0.008, 95% CI [-0.72,-0.11]) discontinuation, indicating consistently declining scores over time. For the naming task, non-carriers had negative trajectories post-discontinuation (*p*=0.021, 95% CI [-0.35,-0.03]), whereas carriers had negative trajectories pre-discontinuation (*p*=0.021, 95% CI [-0.51,-0.04]). Finally, both non-carriers (*p*<.001, 95% CI [2.62,7.31]) and carriers (p=.013, 95% CI [0.85,7.02]) showed positive slopes post-discontinuation for the Trail Making Task B, indicating slower performance following discontinuation, a significant change in trajectory for non-carriers (p=.048, 95% CI [0.04,7.20]).

### Neuroimaging

#### Associations between estradiol use and brain structure differed by APOEε4 status

Cortical thickness, subcortical volumes and white matter hypointensities (WMH) were predicted in a subset of participants with available neuroimaging data, either using E2 or not using MHT (Figure 4A; demographic characteristics in Table 4).

**Figure 4.**
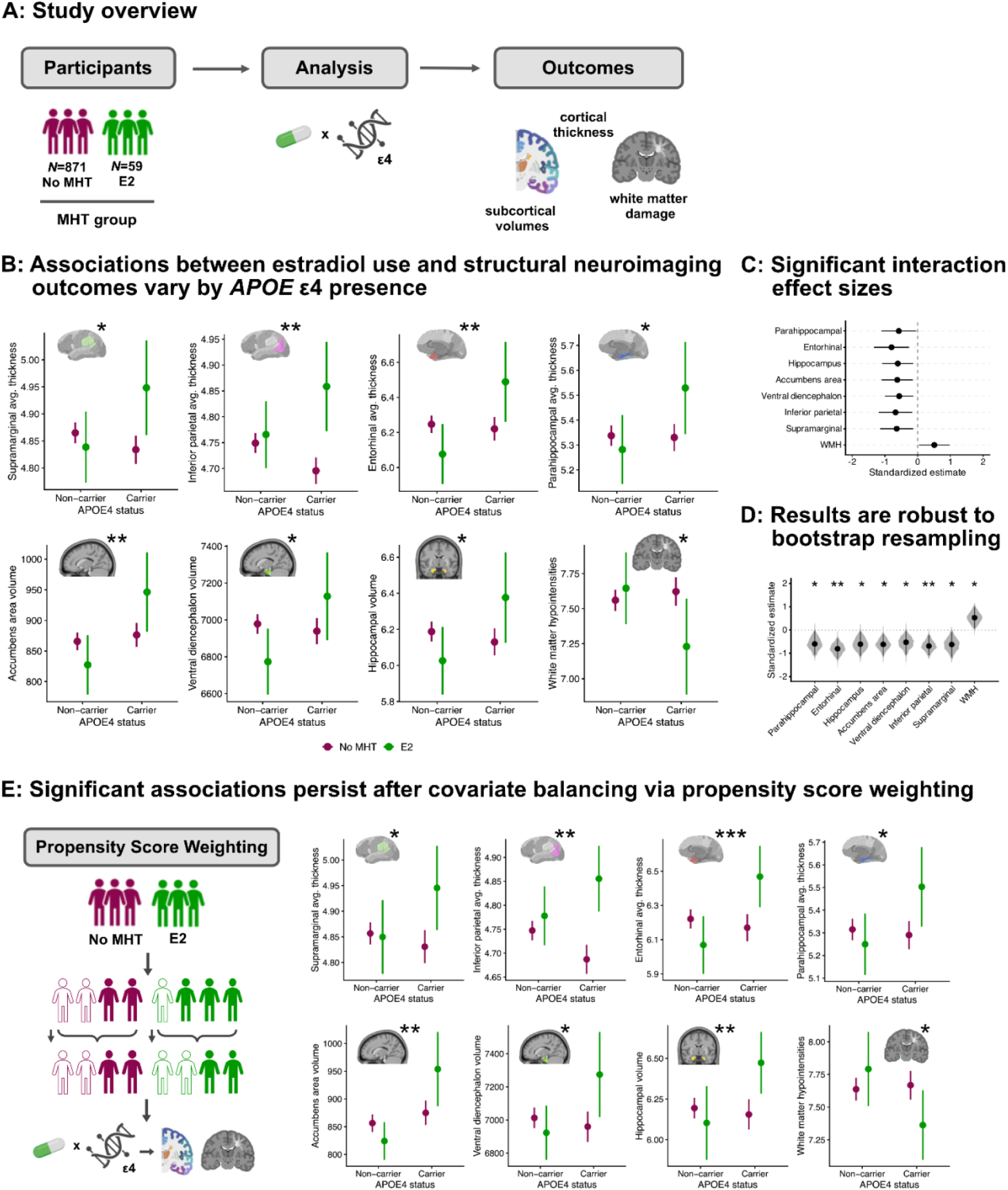
Associations between estradiol therapy use and structural neuroimaging outcomes vary by *APOEε4* status. A: Neuroimaging outcomes were predicted by an interaction between estradiol (E2) therapy use and *APOEε4* status in a subset of participants with available imaging data (no MHT = 871; E2 = 59). B: Significant interactions across brain regions indicated more beneficial associations with E2 use in *APOEε4* carriers than non-carriers (higher cortical thickness and subcortical volumes; lower white matter damage). Models were adjusted for age, years of education, cardiovascular risk, race/ethnicity, and intracranial volume. Plots show model estimates with error bars denoting 95% confidence intervals. C: Standardized effect sizes of significant effects, with values expressed in standard deviation units. D: Results of 1,000 bootstrap resamples per outcome suggest that all significant interaction effects were robust to resampling with replacement. Points represent bootstrapped standardized effect sizes with 95% bootstrap confidence intervals. Grey violins represent the bootstrap sampling distribution of the effect estimate across bootstraps. D: Primary analyses were followed by propensity score weighted sensitivity analyses which showed convergent results for all regions of interest. All significance indicators refer to interaction effects. “*” indicates p < .05, “**” indicates p < .01, “***” indicates p < .001. Images in the first four and the last panel were created with BioRender. Images in the first three panels of the second figure row were adapted under a CC BY 4.0 license, credit: https://doi.org/10.6084/m9.figshare.10063016.v1. The analysis overview figure was created using BioRender.

**Table 4.** Neuroimaging analyses cohort characteristics.

|  | No MHT<br>*n* = 871 | E2<br>*n* = 59 | p-value |
| --- | --- | --- | --- |
| **Age** | 71.2 (7.3) | 70.4 (7.5) | .413 |
| **Years of education** | 16.2 (2.5) | 16.8 (2.1) | .060 |
| **Depressive symptoms** | 1.3 (1.9) | 1.4 (1.9) | .930 |
| **Cardiovascular risk score** | 0.06 (0.14) | 0.10 (0.19) | .049 |
| ***APOE ε4* carriers** | 31.8% | 35.6% | .646 |
| **Race/ethnicity** |  |  |  |
| *American Indian or Alaska Native* | 1.0% | / |  |
| *Asian* | 1.5% | 1.7% |  |
| *Black or African American* | 20.3% | 8.5% |  |
| *Hispanic/Latino* | 10.2% | 6.8% |  |
| *White* | 66.1% | 83.1% |  |
| *Other* | 0.7% | / |  |

NB. Values indicate M (SD) unless % is specified. Race/ethnicity categories reflect wording used in the NACC. Where Hispanic/Latino is not specified, rows reflect percentages for non-Hispanic participants.

Results (Figure 4B) indicated significant interactions between E2 use and *APOEε4* status in predicting average thickness of inferior parietal (*p*=0.009, 95% CI [0.04,0.26]), supramarginal (*p*=0.014, 95% CI [0.32,0.25]), parahippocampal (*p*=0.035, 95% CI [0.02,0.49]), and entorhinal areas (*p*=0.003, 95% CI [0.15,0.73]); as well as accumbens area (*p*=0.011, 95% CI [25.31,191.54]), ventral diencephalon (*p*=0.012, 95% CI [88.35,699.92]), and hippocampal volumes (*p*=0.013, 95% CI [0.09,0.73]); and WMH (*p*=0.032, 95% CI [-0.917,-0.040]). Interaction effects showed the same pattern across outcomes of interest, with more favourable outcomes (higher thickness and volume; lower WMH) seen in *APOEε4* carriers taking E2, but not in non-carriers. All significant interactions were robust to bootstrap resampling (p_boot_<0.05; N_boot_=1,000; (Figure 4D).

The primary analyses were followed by propensity score weighted sensitivity analyses (Figure 4E). Significant interactions persisted following covariate balancing (inferior parietal: *p*=0.004, 95% CI [0.04, 0.23]; supramarginal: *p*=0.032, 95% CI [0.01, 0.23]; parahippocampal: *p*=0.014, 95% CI [0.06, 0.50]; entorhinal: *p*<0.001, 95% CI [0.19, 0.71]; accumbens area: *p*=0.004, 95% CI [35.93, 186.35]; ventral diencephalon: *p*=0.011, 95% CI [90.81, 718.34]; hippocampal: *p*=0.008, 95% CI [0.11, 0.71]; WMH: *p*=0.022, 95% CI [-0.851, -0.068].

## DISCUSSION

The current study examined the associations between MHT formulation and *APOEε4* genotype with cognition in an analysis directly comparing four of the most commonly studied^24^ formulations of MHT. Results indicate that associations between MHT and cognitive performance are formulation- and domain-specific, with E2 showing benefits for episodic memory and both E2 and CEE showing benefits for verbal performance. Critically, beneficial associations between CEE and verbal performance were limited to *APOEε4* non-carriers while E2 also showed beneficial associations in the high-risk *APOEε4* carriers. Evidence of potential benefits of E2 for *APOEε4* carriers extend to brain structure, including the degree of white matter damage.

Results showing that episodic memory was worse in CEE users relative to those not using MHT or using E2 at baseline is in line with prior work showing worse verbal memory performance in CEE users relative to non-users of MHT^44^ as well as a notable literature demonstrating benefits of E2 for the structure and function of the hippocampus^11,15^, the brain’s episodic memory hub. Performance on hippocampal-dependent tasks commonly increases with E2 levels across age groups and hormonal transitions^19,45,46^ in humans. In rodent menopause models, E2 dose-dependently increases, whereas E1 decreases hippocampal-based associative learning and neurogenesis^17,47^. However, while the results of our cross-sectional analyses directly comparing E2 and CEE users consistently suggested a relative benefit of E2, longitudinal analyses showing a decrease in memory scores following CEE discontinuation suggest that CEE may nevertheless contribute to memory performance maintenance to some extent. Similarly, CEE use seemed to help maintain executive function performance in longitudinal analyses. Differential potency between E1-based CEE and E2 may help explain these differences in associations by cognitive domain, as estrogenic effects on cognition vary in a dose-dependent manner^15,23^.

On the other hand, while E2 and CEE both showed positive associations with verbal performance, patterns of associations varied by *APOEε4* status. Both propensity weighted baseline and longitudinal pre/post-discontinuation analyses suggested that positive associations with E2 use were either stronger in *APOEε4* carriers or present for both carriers and non-carriers. In contrast, beneficial associations between CEE and performance on both category fluency and confrontation naming were limited to non-carriers, with longitudinal analyses indicating negative associations between CEE use and naming task performance in *APOEε4* carriers. These findings help clarify prior contradictions in the literature regarding *APOEε4* modulation of MHT effects. Namely, earlier behavioral work^48,49^, with samples comprised predominantly of CEE users, has indicated that MHT use was associated with worse cognitive outcomes in *APOEε4* carriers relative to non-carriers, leading to recommendations to avoid MHT use in *APOEε4* carriers^50^. On the other hand, more recent data^37^, likely collected from majority E2 users, suggested cognitive benefits of MHT use in *APOEε4* carriers. Likewise, E2 use has been associated with less neuropathology in neuroimaging work, with effects particularly pronounced in *APOEε4* carriers^51^. Collectively, these findings demonstrate the importance of differentiating between MHT types, and suggest that modulatory effects of *APOE* genotype may vary by MHT formulation, an idea supported by data from the current study.

Notably, our exploratory analyses indicated that positive associations between MHT and cognition may be particularly pronounced in Black women. Black women using E2 had better scores on verbal and executive function measures relative to those not using MHT, and outperformed those using CEE on episodic memory. This consistent benefit of E2 therapy, including where no significant associations were seen in the overall cohort, provides strong impetus for further research into MHT and Black women’s cognitive health. Critically, Black women in the United States experience earlier onset of menopause and more severe menopause symptoms^41,52^. At the same time, they are significantly less likely to use MHT than white American women^52,53^, despite more severe experiences of vasomotor symptoms and vaginal dryness^52,53^, for which MHT is the first line of treatment. Scholarship in Black Feminist-Womanist Gerontology has identified multiple compounding reasons for these disparities including institutional barriers, socioeconomic and cultural factors, and healthcare access^54,55^. Yet despite these well-documented disparities, Black women are underrepresented in the literature on MHT-related outcomes^56^.

As prior research, albeit in largely white cohorts, has found that higher menopause symptom burden is associated with worse cognitive outcomes^9^, it is not surprising that higher benefits of MHT would be seen in a group statistically more likely to have experienced more severe menopause symptoms, including vasomotor ones, the negative effects of which have been particularly well-documented in the cognitive aging literature^9^. Although the current findings are exploratory, they demonstrate the importance of considering disparities in menopause care and their effects on cognitive aging, with potential implications for menopause care in Black women. Additional research is needed to replicate these effects and elucidate their mechanism, including structural barriers to menopause care access, in larger samples with higher availability of both reproductive history data and sociodemographic information beyond simple race categories^57,58^.

The fact that we primarily identified positive associations with cognition in unopposed estrogen groups is consistent with prior observational studies of MHT. For example, a recent meta-analysis showed reduced risk of all-cause dementia associated with unopposed estrogens, but not with a combination MHT^22^. Route of administration is important to consider when interpreting effects of unopposed estrogens. While orally administered unopposed estrogens are only clinically recommended post-hysterectomy (although pharmacy claims data suggest these guidelines are not always followed^59,60^), unopposed formulations also come in transdermal and vaginal forms, with concurrent progesterone use not being required for the latter. The latter consideration is particularly relevant as vaginal (and therefore unopposed) formulations tend to be more common among older adults in the age range of participants in the current study^61–63^. It is also possible that some used levonorgestrel intrauterine devices for endometrial protection which may have gone underreported^28^. However, as data on route of administration and gynecological surgery history in the NACC was lacking, it was not possible for us to directly compare these MHT user subgroups. We encourage the addition of this information to longitudinal cohort studies.

In addition to significant associations between MHT and cognitive performance, neuroimaging analyses indicated a consistent pattern of more favourable outcomes in *APOEε4* carriers using E2, including higher cortical thickness, subcortical volumes, and fewer white matter lesions. These findings replicate prior work indicating that estrogen-based MHT use is associated with fewer white matter lesions^37^ and higher hippocampal volume^37,38^, with particularly strong effects in *APOEε4* carriers. Interestingly, emerging work taking MHT route of administration into account indicates that topical applications are associated with fewer WMH relative to oral MHT (unspecified type)^64^. In line with findings on WMH and hippocampal volume, prior neuroimaging work examining AD pathology in E2 users specifically found more pronounced positive effects in APOEε4 carriers^51,65^. The current results also demonstrate positive associations between E2 use in *APOEε4* carriers and cortical thickness of parietal regions as well as volumes of subcortical regions involved in the reward circuitry, suggesting that MHT use may be associated with structural differences across a broader range of regions beyond the medial temporal lobe.

Although parietal regions and subcortical regions associated with reward processing have been underexplored in the context of MHT, connectivity of these areas has been associated with E2 levels during the female reproductive lifespan^66–68^. These regions also matter for aging and AD risk. Parietal degeneration is particularly predictive of early AD^69–72^, and sex difference analyses of *APOEε4* effects on functional connectivity in older, cognitively healthy adults indicate higher vulnerability to reduced parietal connectivity in female, but not male, *APOEε4* carriers^35^. The current findings suggest that E2 use may buffer against this risk in female *APOEε4* carriers. Similarly, nucleus accumbens shows more AD neuropathology relative to other striatal regions^73,74^ and its lower volume is predictive of AD^75,76^. Our results indicate potential protective effects of E2 for this region in *APOEε4* carriers. Collectively, the current results indicate positive associations between E2 therapy and structural outcomes in *APOEε4* carriers across brain regions implicated in early AD.

Key strengths of the current study include precise characterization of MHT formulation, inclusion of both cognitive performance and neuroimaging data, characterization of both current use and post-discontinuation effects of MHT, and direct comparison of different types of MHT formulations. While MHTs based on different types of estrogens have been examined in isolation, few studies have directly compared different types of estrogen-based MHTs to a non-user group (a notable exception is the KEEPs trial^77,78^) and, to our knowledge, no prior studies have directly compared unopposed and combined formulations of different estrogen-based MHTs in one design including a comparison group not taking MHT. Additionally, propensity score weighted sensitivity analyses helped reduce the impact of potential selection bias among participants taking MHT. Key limitations of the current data include lack of information on the route of MHT administration, which matters for both cognitive^25^ and brain^64^ aging, as well as dose and length of use. Future work would benefit from larger cohorts, more detailed reproductive history data, including past reproductive experiences such as parity, age and type of menopause (surgical vs. spontaneous), as well as information on MHT use duration, age at initiation, and route of administration. Lack of detailed reproductive history information in the current data limited the extent to which we could adjust for baseline characteristics using propensity score weighting. Finally, it is important to note that our cross-sectional neuroimaging results could only speak to associations of current MHT use with structural brain outcomes, which may not generalize to past users^79^. Patterns of past use and their effects on brain health should be considered in future work.

The current results suggest that MHT effects on cognition are formulation- and domain-specific, with evidence of E2 being more beneficial for *APOEε4* carriers, who are at increased risk of AD. Our results highlight the importance of considering MHT formulation and presence of individual risk factors when assessing cognitive impact of MHT, reframe mixed MHT findings as an issue of exposure definition, and support hormonal composition- and genetic-risk-aware approaches to menopause care.

## Data Availability

All data are available NAcc. https://www.naccdata.org/ Accessing the data involves submitting a formal data request and adherence to the NACC data use agreement.

## METHODS

### Participants

Participant data was obtained from the National Alzheimer’s Coordinating Center (NACC^1^). NACC recruits from over 40 Alzheimer’s and dementia research centers across the United States, and collects clinical, cognitive, neuroimaging, and genotype data. The inclusion criteria for the current study were: cognitively healthy individuals (defined by a Clinical Dementia Rating scale ^2^ score of 0), aged 50-90, without traumatic brain injury or cancer diagnoses. We used baseline visit NACC data for the primary cognitive analyses; longitudinal data from participants with at least two consecutive visits where use of MHT was reported followed by at least two consecutive visits without reported MHT use for the pre-/post-discontinuation secondary analyses; and MRI data collected within ± 6 months of a clinical visit for the neuroimaging analyses. MHT formulation information was extracted from participant medication records.

After exclusions, the primary cognitive analysis sample included 8,741 participants (no MHT = 7,955; E2 = 414; E2+P = 65; CEE = 243; CEE+MPA = 64). Thirteen participants in the E2+P group used a progestin combination (estradiol-norethindrone), the rest used E2 combined with progesterone. Route of MHT administration was not consistently reported. Most medication fields only specified hormonal formulation; however, “topical” was flagged for 21.3% of participants using E2 and 6.17% of participants using CEE. The pre-/post-discontinuation sample included 165 participants (N_E2_ = 111; N_CEE_ = 54) with an average of 3.3 ± 2 visits available pre-discontinuation and an average of 4.6 ± 2.9 visits available post-discontinuation. The neuroimaging analysis sample included 930 participants with available imaging data (no MHT = 871; E2 = 59).

### Cognitive Tasks

Cognitive outcomes of interest were episodic memory, verbal function (category fluency, confrontation naming), and executive function – domains previously shown to vary across the menopausal transition and as a function of MHT use^3–5^. The NACC neuropsychological battery was revised in 2015 with multiple tests replaced. Subsequently NACC released a validated conversion strategy for harmonizing the implicated tests with the new versions^6^. We used these guidelines to harmonize results from the initial Logical Memory and the later Craft Story^7^, which measure episodic memory, and the initial Boston Naming Test and the later Multilingual Naming Test^8,9^, which measure confrontation naming. Additionally, category fluency^10^ (computed as the average between animal and vegetable categories) and Trail Making Test B^11^ data were available for all participants and used in their original form. The latter was used as a measure of executive function.

### Neuroimaging

Cortical thickness, subcortical volumes and white matter hypointensities (WMH) were predicted in a subset of participants with neuroimaging data. Extracted structural data was obtained from the NACC’s Standardized Centralized Alzheimer’s and Related Dementias Neuroimaging (SCAN) dataset. SCAN protocols are summarized in publicly available NACC documentation^12^. Primary regions of interest for the current analyses involved medial temporal lobe regions which are among the first to be affected by AD pathology (hippocampus, entorhinal cortex, parahippocampal cortex)^13^. WMH (log transformed) were also a primary outcome of interest due to associations with both MHT use^14^ and AD risk^15^. Additionally, while interaction effects between *APOE* genotype and MHT use on temporal regions and a notable portion of the reward circuit (prefrontal cortex, anterior cingulate, and the amygdala) have been explored in past work^14,16^, their effects on other reward system regions and the parietal cortex remain unexamined. These are important gaps as these regions are both sensitive to estradiol and implicated in AD^17–22^. We thus included four parietal (superior and inferior parietal, supramarginal gyrus, precuneus) and five subcortical reward regions (accumbens area, dorsal striatum, ventral diencephalon, thalamus, pallidum) in the analysis as exploratory regions of interest.

### Additional measures

Cardiovascular risk and depressive symptoms were considered as potential confounding variables. Depressive symptoms were measured with the Geriatric Depression Scale ^23^. Scores ranged from 0-30, based on yes/no responses to 30 items measuring depressive symptoms (e.g., “Do you often feel helpless?”). Higher scores indicated higher likelihood of depression (0-9: low likelihood of depression; 10-19: likely mild depression; 20-30: likely severe depression). Cardiovascular risk was estimated using an established composite score calculation which has previously been used with NACC data ^24–26^, based on the average of the following dichotomous variables: hypertension, hypercholesterolemia, diabetes, stroke, and heart conditions (heart attack, congestive heart failure, cardiac bypass surgery, and/or angina). Total scores ranged from 0-1 with higher scores indicating more risk.

### Statistical analysis

#### Primary analyses

All analyses were conducted in R Studio (R version 4.5.1). Primary cognitive and neuroimaging outcomes were predicted by an interaction between group (no MHT, E2, E2+P4, CE, CE+MPA) and *APOEε4* presence using linear models. Age, education, and race/ethnicity were included as covariates in all analyses. Additional modifiable risk factors (depressive symptoms, cardiovascular risk score) were included as covariates if significant group differences were present. Consequently, age, race/ethnicity, depressive symptoms, and years of education were added as covariates in cognitive analyses, and age, race/ethnicity, education and cardiovascular risk were included as covariates in neuroimaging analyses. Additionally, total intracranial volume was included as a covariate in neuroimaging analyses. Robustness of observed effects was examined through nonparametric bootstrap resampling with replacement (N_boot_ = 1,000).

Primary analyses were followed by propensity score weighted sensitivity analyses to improve covariate balance, accounting for potential selection bias in the MHT groups. Propensity score weighing was conducted using the WeightIt package in R^27^, specialized for estimating balancing weights for observational studies. Weights were estimated with the covariate balancing propensity score and targeted the average treatment effect in the overlap population. The propensity score model included age, education, race/ethnicity, depressive symptoms and cardiovascular risk, with estimates by *APOEε4* status. To further reduce potential residual confounding, weighted analyses included covariate adjustment mirroring the primary analytic models for a doubly-adjusted approach. The propensity score weighted sensitivity analysis for the cognitive outcomes involved a reduced sample (no MHT, E2, CEE) as initial weighting of the full analytic sample yielded very low effective sample sizes for the combination therapy groups. Propensity score weighting for both cognitive (effective sample sizes for no MHT = 6,510; E2 = 352; CEE = 224) and neuroimaging outcomes (effective sample sizes for no MHT = 602; E2 = 59) demonstrated successful covariate balance (all absolute standardized mean differences < 0.1).

#### Secondary analyses

As a complement to the primary cross-sectional analyses, trajectories before and after MHT discontinuation were analyzed using piecewise linear mixed-effect models in a subset of participants with at least two consecutive visits during which MHT use was reported (pre-discontinuation) followed by at least two consecutive visits during which no MHT use was reported (post-discontinuation). Discontinuation was estimated at the midpoint between the pre- and post-discontinuation visits. Models included interactions between *APOEε4* and the pre- and post-discontinuation visit segments, random intercepts for participants, and random slopes for time. All models were adjusted for age at baseline, race/ethnicity, depressive symptoms, cardiovascular risk, years of education, and the cumulative proportion of visits for which participants had reported MHT use over the course of NACC data collection. Square root of visit number was added as a covariate to models predicting episodic memory and naming task performance as visual inspection suggested presence of practice effects (Extended Figure 2). Linear contrasts were used to estimate the difference between the post- and pre-discontinuation slopes as a measure of the degree of trajectory change following MHT discontinuation. Modeling was performed separately in the E2 (N = 111) and CEE (N = 54) user samples, as we were interested in within-subject change following discontinuation.

#### Exploratory analyses

Exploratory analyses examined cognitive outcomes as a function of menopause hormone therapy use and race. Due to sample size constraints, only non-Hispanic Black and white participants who either did not take MHT (N = 6,997) or took E2 (N = 383) or CEE (N = 235) were included in the analyses, and *APOEε4* presence was included as a covariate, not an interaction term. Cognitive performance was thus predicted by an interaction between MHT group (no MHT, E2, CEE) and race using linear models adjusted for age, cardiovascular risk, years of education and *APOEε4*. As cognitive outcomes associated with MHT use in Black participants were of primary interest, all analyses were conducted with Black participants as the reference group.

## Acknowledgements

Creation of all figures involved use of BioRender.com. Citation: Created in BioRender. Galea, L. (2026) https://BioRender.com/kxlkwtu

## Author Contributions

MP and LAMG conceptualized the study. MP designed and conducted analyses, wrote the manuscript, and produced the figures. MWA aided in figure design. LAMG supervised the study and provided resources. LAMG, JSR, MP, AJMG, and LLG were responsible for funding acquisition for the study. All authors provided conceptual contributions and reviewed and edited the final manuscript.

## Funding statement

This project was supported by a CIHR Postdoctoral Fellowship (MFE-201012) to M.P., Wellcome Leap Inc funding (CARE-2025-2571407103) to L.A.M.G., J.S.R., A.J.MG., M.P. and L.L.G., womenmind^TM^ (CAMHF-1197) to L.A.M.G, Alzheimer Society of Canada Research Program to M.W.A. and J.S.R., CIHR (173253, 438475) to J.S.R., Canada Graduate Scholarships program to M.W.A., The Alzheimer’s Association to J.S.R. The NACC database is funded by NIA/NIH Grant U24 AG072122. NACC data are contributed by the NIA-funded ADRCs: P30 AG062429 (PI James Brewer, MD, PhD), P30 AG066468 (PI Oscar Lopez, MD), P30 AG062421(PI Bradley Hyman, MD, PhD), P30 AG066509 (PI Thomas Grabowski, MD), P30 AG066514 (PI Mary Sano, PhD), P30 AG066530 (PI Helena Chui, MD), P30 AG066507 (PI Marilyn Albert, PhD), P30 AG066444 (PI John Morris, MD), P30 AG066518 (PI Jeffrey Kaye, MD), P30 AG066512 (PI Thomas Wisniewski, MD), P30 AG066462 (PI Scott Small, MD), P30 AG072979 (PI David Wolk, MD), P30 AG072972 (PI Charles DeCarli, MD), P30 AG072976 (PI Andrew Saykin, PsyD), P30 AG072975 (PI David Bennett, MD), P30 AG072978 (PI Neil Kowall, MD), P30 AG072977 (PI Robert Vassar, PhD), P30 AG066519 (PI Frank LaFerla, PhD), P30 AG062677 (PI Ronald Petersen, MD, PhD), P30 AG079280 (PI Eric Reiman, MD), P30 AG062422 (PI Gil Rabinovici, MD), P30 AG066511 (PI Allan Levey, MD, PhD), P30 AG072946 (PI Linda Van Eldik, PhD), P30 AG062715 (PI Sanjay Asthana, MD, FRCP), P30 AG072973 (PI Russell Swerdlow, MD), P30 AG066506 (PI Todd Golde, MD, PhD), P30 AG066508 (PI Stephen Strittmatter, MD, PhD), P30 AG066515 (PI Victor Henderson, MD, MS), P30 AG072947 (PI Suzanne Craft, PhD), P30 AG072931 (PI Henry Paulson, MD, PhD), P30 AG066546 (PI Sudha Seshadri, MD), P20 AG068024 (PI Erik Roberson, MD, PhD), P20 AG068053 (PI Justin Miller, PhD), P20 AG068077 (PI Gary Rosenberg, MD), P20 AG068082 (PI Angela Jefferson, PhD), P30 AG072958 (PI Heather Whitson, MD), P30 AG072959 (PI James Leverenz, MD). The NACC database is funded by NIA/NIH Grant U24 AG072122. SCAN is a multi-institutional project that was funded as a U24 grant (AG067418) by the National Institute on Aging in May 2020. Data collected by SCAN and shared by NACC are contributed by the NIA-funded ADRCs as follows: Arizona Alzheimer’s Center - P30 AG072980 (PI: Eric Reiman, MD); R01 AG069453 (PI: Eric Reiman (contact), MD); P30 AG019610 (PI: Eric Reiman, MD); and the State of Arizona which provided additional funding supporting our center; Boston University - P30 AG013846 (PI Neil Kowall MD); Cleveland ADRC - P30 AG062428 (James Leverenz, MD); Cleveland Clinic, Las Vegas - P20AG068053; Columbia - P50 AG008702 (PI Scott Small MD); Duke/UNC ADRC - P30 AG072958; Emory University - P30AG066511 (PI Levey Allan, MD, PhD); Indiana University - R01 AG19771 (PI Andrew Saykin, PsyD); P30 AG10133 (PI Andrew Saykin, PsyD); P30 AG072976 (PI Andrew Saykin, PsyD); R01 AG061788 (PI Shannon Risacher, PhD); R01 AG053993 (PI Yu-Chien Wu, MD, PhD); U01 AG057195 (PI Liana Apostolova, MD); U19 AG063911 (PI Bradley Boeve, MD); and the Indiana University Department of Radiology and Imaging Sciences; Johns Hopkins - P30 AG066507 (PI Marilyn Albert, Phd.); Mayo Clinic - P50 AG016574 (PI Ronald Petersen MD PhD); Mount Sinai - P30 AG066514 (PI Mary Sano, PhD); R01 AG054110 (PI Trey Hedden, PhD); R01 AG053509 (PI Trey Hedden, PhD); New York University - P30AG066512-01S2 (PI Thomas Wisniewski, MD); R01AG056031 (PI Ricardo Osorio, MD); R01AG056531 (PIs Ricardo Osorio, MD; Girardin Jean-Louis, PhD); Northwestern University - P30 AG013854 (PI Robert Vassar PhD); R01 AG045571 (PI Emily Rogalski, PhD); R56 AG045571, (PI Emily Rogalski, PhD); R01 AG067781, (PI Emily Rogalski, PhD); U19 AG073153, (PI Emily Rogalski, PhD); R01 DC008552, (M.-Marsel Mesulam, MD); R01 AG077444, (PIs M.-Marsel Mesulam, MD, Emily Rogalski, PhD); R01 NS075075 (PI Emily Rogalski, PhD); R01 AG056258 (PI Emily Rogalski, PhD); Oregon Health and Science University - P30 AG008017 (PI Jeffrey Kaye MD); R56 AG074321 (PI Jeffrey Kaye, MD); Rush University - P30 AG010161 (PI David Bennett MD); Stanford - P30AG066515; P50 AG047366 (PI Victor Henderson MD MS); University of Alabama, Birmingham - P20; University of California, Davis - P30 AG10129 (PI Charles DeCarli, MD); P30 AG072972 (PI Charles DeCarli, MD); University of California, Irvine - P50 AG016573 (PI Frank LaFerla PhD); University of California, San Diego - P30AG062429 (PI James Brewer, MD, PhD); University of California, San Francisco - P30 AG062422 (Rabinovici, Gil D., MD); University of Kansas - P30 AG035982 (Russell Swerdlow, MD); University of Kentucky - P30 AG028283-15S1 (PIs Linda Van Eldik, PhD and Brian Gold, PhD); University of Michigan ADRC - P30AG053760 (PI Henry Paulson, MD, PhD) P30AG072931 (PI Henry Paulson, MD, PhD) Cure Alzheimer’s Fund 200775 - (PI Henry Paulson, MD, PhD) U19 NS120384 (PI Charles DeCarli, MD, University of Michigan Site PI Henry Paulson, MD, PhD) R01 AG068338 (MPI Bruno Giordani, PhD, Carol Persad, PhD, Yi Murphey, PhD) S10OD026738-01 (PI Douglas Noll, PhD) R01 AG058724 (PI Benjamin Hampstead, PhD) R35 AG072262 (PI Benjamin Hampstead, PhD) W81XWH2110743 (PI Benjamin Hampstead, PhD) R01 AG073235 (PI Nancy Chiaravalloti, University of Michigan Site PI Benjamin Hampstead, PhD) 1I01RX001534 (PI Benjamin Hampstead, PhD) IRX001381 (PI Benjamin Hampstead, PhD); University of New Mexico - P20 AG068077 (Gary Rosenberg, MD); University of Pennsylvania - State of PA project 2019NF4100087335 (PI David Wolk, MD); Rooney Family Research Fund (PI David Wolk, MD); R01 AG055005 (PI David Wolk, MD); University of Pittsburgh - P50 AG005133 (PI Oscar Lopez MD); University of Southern California - P50 AG005142 (PI Helena Chui MD); University of Washington - P50 AG005136 (PI Thomas Grabowski MD); University of Wisconsin - P50 AG033514 (PI Sanjay Asthana MD FRCP); Vanderbilt University - P20 AG068082; Wake Forest - P30AG072947 (PI Suzanne Craft, PhD); Washington University, St. Louis - P01 AG03991 (PI John Morris MD); P01 AG026276 (PI John Morris MD); P20 MH071616 (PI Dan Marcus); P30 AG066444 (PI John Morris MD); P30 NS098577 (PI Dan Marcus); R01 AG021910 (PI Randy Buckner); R01 AG043434 (PI Catherine Roe); R01 EB009352 (PI Dan Marcus); UL1 TR000448 (PI Brad Evanoff); U24 RR021382 (PI Bruce Rosen); Avid Radiopharmaceuticals / Eli Lilly; Yale - P50 AG047270 (PI Stephen Strittmatter MD PhD); R01AG052560 (MPI: Christopher van Dyck, MD; Richard Carson, PhD); R01AG062276 (PI: Christopher van Dyck, MD); 1Florida - P30AG066506-03 (PI Glenn Smith, PhD); P50 AG047266 (PI Todd Golde MD PhD)

## Competing interests

All authors declare they have no competing interests.

## Data availability statement

NACC data can be accessed through the following website: https://www.naccdata.org/ Accessing the data involves submitting a formal data request and adherence to the NACC data use agreement.

## EXTENDED FIGURES

**Extended Figure 1.**
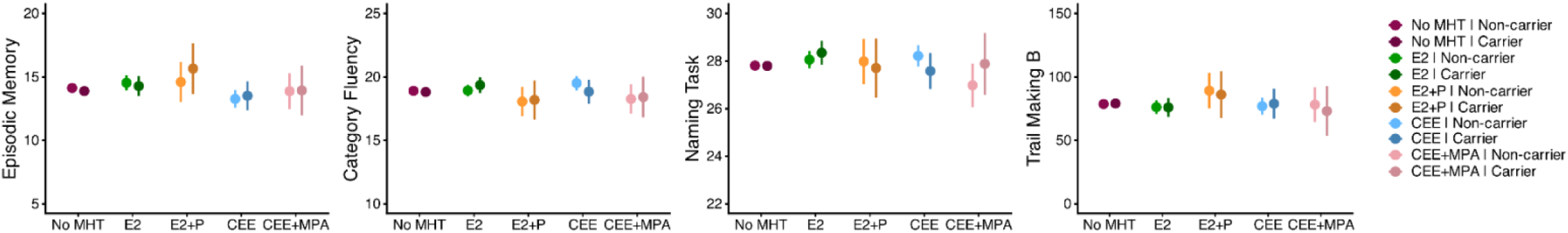
Cognitive effects of menopause hormone therapy broken down by APOE status.

**Extended Figure 2.**
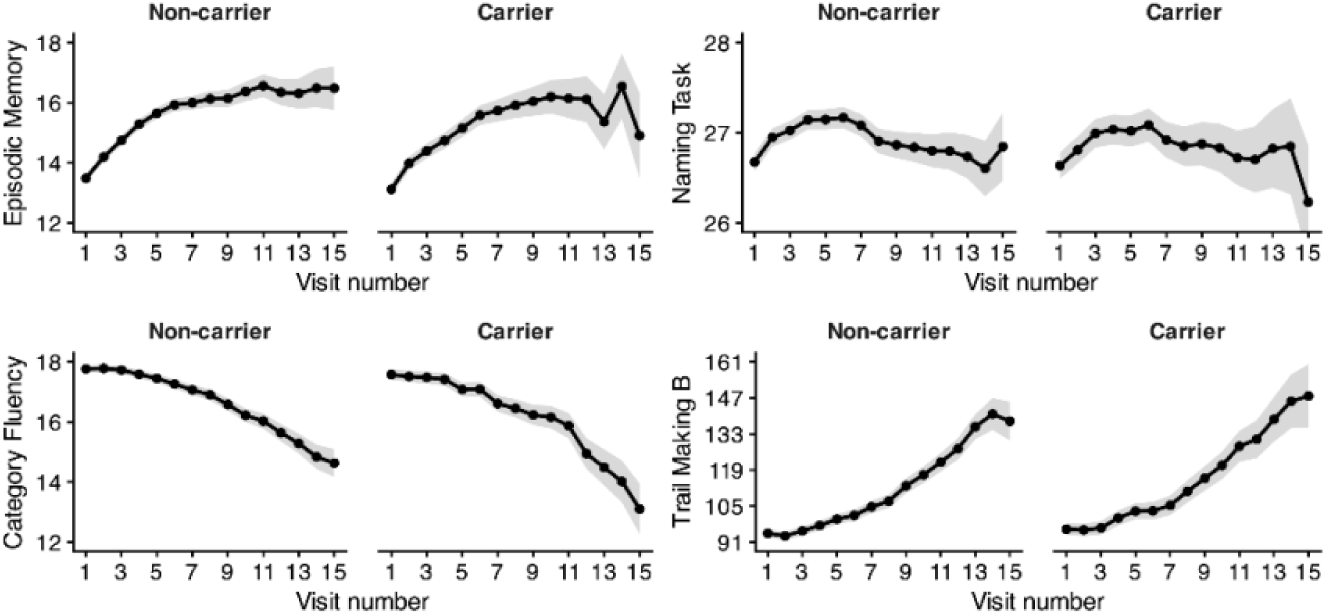
Age-adjusted cognitive performance curves in a comparison group of participants not on MHT indicate presence of practice effects for episodic memory and the naming task.

## Notes

### Competing Interest Statement

The authors have declared no competing interest.

### Author Declarations

This study used data available upon request from https://www.naccdata.org/

